# Seroprevalence of SARS-CoV-2 IgG antibodies in two regions of Estonia (KoroSero-EST-1)

**DOI:** 10.1101/2020.10.21.20216820

**Authors:** Piia Jõgi, Hiie Soeorg, Diana Ingerainen, Mari Soots, Freddy Lättekivi, Paul Naaber, Karolin Toompere, Pärt Peterson, Liis Haljasmägi, Eva Žusinaite, Hannes Vaas, Merit Pauskar, Arina Shablinskaja, Katrin Kaarna, Heli Paluste, Kai Kisand, Marje Oona, Riina Janno, Irja Lutsar

## Abstract

**Background:** In Estonia, during the first wave of COVID-19 total number of cases confirmed by PCR was 13.3/10,000, similar in most regions, including capital Tallinn, but in the hotspot of Estonian epidemic, an island Saaremaa, the cumulative incidence was 166.1/10,000.

**Aim:** We aimed to determine the prevalence of SARS-CoV-2 IgG antibodies in these two regions, symptoms associated with infection and factors associated with antibody concentrations.

**Methods:** Participants were selected using stratified (formed by age decades) random sampling and recruited by general practitioners. IgG were determined from sera by four assays. Symptoms of acute respiratory illness associated with seropositivity were analyzed by multiple correspondence analysis, antibody concentrations by multiple linear regression.

**Results:** Total of 3608 individual were invited and 1960 recruited From May 8 to July 31, 2020. Seroprevalence was 1.5% (95% confidence interval (CI) 0.9-2.5) and 6.3% (95% CI 5.0-7.9), infection fatality rate 0.1% (95% CI 0.0-0.2) and 1.3% (95% CI 0.4-2.1) in Tallinn and Saaremaa, respectively. Of seropositive subjects 19.2% (14/73) had acute respiratory illness. Fever, diarrhea and the absence of cough and runny nose were associated with seropositivity in individuals aged 50 or more years. IgG concentrations were higher if fever, difficulty breathing, shortness of breath, chest pain or diarrhea was present, or hospitalization required.

**Conclusion:** Similarly to other European countries the seroprevalence of SARS-CoV-2 in Estonia was low even in the hotspot region Saaremaa suggesting that majority of population is still susceptible to SARS-CoV-2. Focusing only on respiratory symptoms may delay accurate diagnosis of SARS-CoV-2 infection.

## Introduction

The first case of COVID-19 was reported in Estonia on February 26, 2020. Two weeks later extensive spread of SARS-CoV-2 virus occurred that led to lockdown, including closure of educational institutions at all levels, recommendations to stay and work at home whenever possible and limitations to public gatherings from March 12, 2020 [1]. By the end of April, the first wave of COVID-19 had been largely contained by the implemented measures that were subsequently lifted or partially relaxed on May 16, 2020. By then, the total cumulative number of cases confirmed by PCR for SARS-CoV-2 in Estonia was 13.3 per 10,000 inhabitants, similar in most of the regions, including the capital Tallinn located in Harju county [2]. In contrast, in the hotspot of Estonian epidemic, an island Saaremaa, the cumulative incidence was 166.1 cases per 10,000 and with its peak incidence of 954.5 cases per 100,000 within last 14 days it was among the European within-country regions experiencing the most extensive spread [1]. However, as the infection-to-case ratio, i.e. the ratio of the seroprevalence to rate of confirmed cases of COVID-19, can vary widely from 6 to 23 times [3], the actual prevalence of infection with SARS-CoV-2 remains unknown in Estonia. In general, the rate of SARS-CoV-2 seropositivity in previously conducted studies have been low, ranging from 0.4 to 10.9% in population-based seroepidemiological studies [4-8]. To accurately estimate the extent of past spread and potential for future spread, seroepidemiological studies are warranted.

In KoroSero-EST-1 study, we aimed to understand the actual prevalence of the infection by determining the seroprevalence of COVID-19 in individuals from two regions in Estonia with very different cumulative incidence of COVID-19. Second, we aimed to describe the symptoms associated with COVID-19 in seropositive individuals and the factors associated with antibody concentrations against SARS-CoV-2.

## Methods

### Study design

The KoroSero-EST-1 was a cross-sectional seroepidemiological study conducted from May 8 to July 31, 2020 in two general practitioner (GP) practices in capital Tallinn in Harju county and in Saaremaa with total number of patients 13,260 and 7,525, respectively.

Participants were selected using stratified random sampling. Strata were formed by classifying patients from each GP into 10-year age groups, except the age group of 80 years or older due to their small number. From each stratum individuals were randomly identified by Estonian Health Insurance Fund with the aim to include at least 110 participants per age group from both GP practices to achieve desirable precision for the seroprevalence estimates.

Participants filled in a questionnaire based on World Health Organization recommendations [9] that included the presence of acute respiratory illness since March 1, 2020 and if present, its symptoms, known contact with PCR-confirmed COVID-19 cases, previously performed PCR test for SARS-CoV-2 and its result (Supplementary Information).

All participants and/or their legal guardians provided written informed consent. The study was approved by the Research Ethics Committee of the University of Tartu.

### Sampling and antibody measurements

Blood samples (3.5 mL) were drawn and stored for 48 hours at +4°C until transported to laboratory, where sera were stored at -30° C until testing in SYNLAB Estonia Central Laboratory in Tallinn or at the research laboratories of the University of Tartu, Estonia.

First, all samples were tested by chemiluminescent microparticle immunoassay for detection of IgG against SARS-CoV-2 nucleoprotein (N) (Abbott Architect SARS-CoV-2 IgG with ARCHITECT i2000SR analyzer; Abbott Laboratories, USA) according to the manufacturer’s instructions. Samples close to cut-off value (signal/cut-off ratio 0.3 to 1.39) were additionally tested by chemiluminescence immunoassay for detection of IgG against S1 and S2 proteins (LIAISON® SARS-CoV-2 S1/S2 IgG test; DiaSorin S.p.A., Italy) to increase sensitivity without influencing specificity [10].

Next, sera positive for anti-N or anti-S1/S2 IgG or those from participants with previously positive PCR test and two age- and sex-matched negative controls or a random subset of them (Figure 1) were tested for anti-N IgG by a rapid Standard Q COVID-19 IgM/IgG Duo test (SD BIOSENSOR, Republic of Korea) according to the manufacturer’s instructions and by in-house luciferase immunoprecipitation system (LIPS) assays as described elsewhere [11].

**Figure 1.**
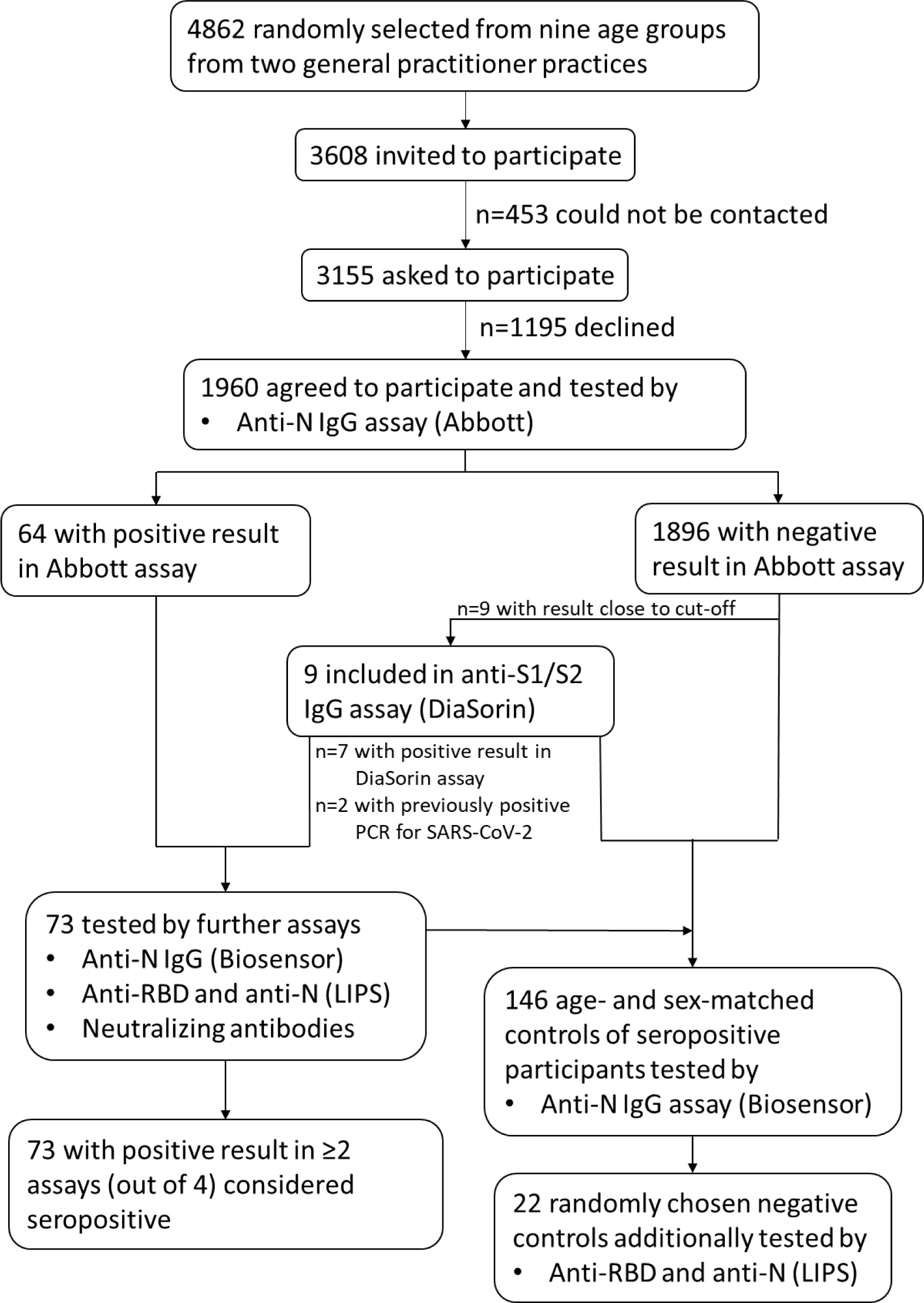
Flowchart of recruitment of study participants, testing and definition for being considered seropositive

Finally, an in-house neutralization assay was performed in duplicates for all PCR-positive samples and samples positive for anti-N or anti-S1/S2 IgG. Briefly, patients’ sera were two-fold serially diluted in Virus Growth Media (VGM – DMEM/0.2% BSA/Pen-Strep) starting from 1:4 to 1:4096 in a volume of 50 μl per well. The viral stock was diluted in VGM to contain 100 pfu (plaque forming units) per 50 μl. 100 plaque forming units of the virus stock per well (a local Estonian SARS-CoV-2 isolate from a COVID-19-positive patient’s nasal swab; from SYNLAB) was added to the wells containing sera dilutions, and the mixtures were incubated for 1 hour at 37°C. After this, 4×10^4^ Vero E6 cell suspension in 100 μl of VGM per well was added to the wells. Plates were incubated at 37°C and 5% CO2 for up to 5 days. Neutralization titer was calculated by microscopically evaluating cytopathic effect (CPE) in infected wells (round/floating cells compared to untouched monolayer in non-infected wells). The dilution of sera that did not show CPE was considered as neutralization titer.

### Statistical analysis

The software program Stata 14.2 was used for calculating seroprevalence estimates and infection fatality rates and R (version 3.6.1) for other statistical analyses.

Total cumulative number of confirmed COVID-19 cases per 10,000 inhabitants was calculated as of two weeks before the midpoint of sample collection in GP practice. Daily numbers of confirmed COVID-19 cases (positive PCR tests for SARS-CoV-2 from nasopharyngeal swabs) in each age group in Harju county (assumed to reflect the spread of SARS-CoV-2 in Tallinn; 72.3% of Harju county inhabitants live in Tallinn) and Saaremaa were obtained from Health Board [2] and the number of inhabitants as of January 1, 2020 from Statistics Estonia.

### Sample size calculation

Sample size of at least 110 participants per stratum was needed to obtain binomial exact 95% CI with width of 5%, assuming seroprevalence was 1% or to estimate seroprevalence with precision ±10%, assuming seroprevalence was more than 10%. Assuming non-response of about 50% (up to 70% in children), 2341 individuals were sampled from both GP practice to whom invitations were sent until about 110 per stratum agreed.

### Seroprevalence estimation

Seroprevalence was estimated as the proportion of participants considered seropositive, i.e. with positive result in at least two different antibody assay out of four assays performed (Figure 1), overall and in each age group, separately for the GP practices. To account for stratification, for each age group in a GP practice sampling weight was used that was further adjusted for response probability by post-stratification. Due to large sampling fractions, finite population correction factor was applied. Binomial exact 95% CI were calculated for seroprevalences in age groups.

Seroprevalence adjusted for test performance of anti-N IgG assay (Abbott Architect SARS-CoV-2 IgG with ARCHITECT i2000SR analyzer) was calculated as

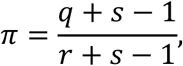

where *r* is sensitivity, *s* specificity and *q* denotes prevalence [12]. Nonparametric bootstrap confidence intervals were calculated for adjusted seroprevalence estimates by drawing 10,000 bootstrap samples at random from our dataset and binomial distribution of the assay validation dataset. Each bootstrap sample was adjusted for sensitivity of 92.7% and specificity of 99.9% [13] and 2.5th and 97.5th percentiles of the bootstrap distributions were reported. Infection fatality rate was calculated as number of reported death cases related to COVID-19 as of July 31, 2020 divided by the estimated total number of seropositive individuals. Confidence intervals were calculated assuming that number of deaths follow Poisson distribution.

### Analysis of symptoms

The association between the presence of symptoms and seropositivity was analyzed by multiple correspondence analysis (MCA) using R package FactoMineR. Only dimensions that explained >1/Q, where Q is the number of variables used, of the total inertia were retained for interpretation. Association between seropositivity and symptoms was determined from dimensions with high contribution and quality (cos^2^>0.1) of seropositivity. Hierarchical clustering using Euclidean distance and Ward’s method followed by k-means consolidation was performed on all dimensions of MCA to partition the individuals into groups. The number of clusters was based on elbow of the barplot of the gain in within-inertia.

For comparing categorical variables, Fisher’s exact test was used.

### Analysis of antibody concentrations/titer

Multiple linear regression was used to analyze the association between IgG and neutralizing (log-transformed and undetectable titers equaled to 0) antibodies concentrations/titer and the presence of acute respiratory illness or its symptoms or hospitalization adjusted for age, sex and time since onset of symptoms, shown to influence antibody concentrations [5, 14].

Multiple imputation was used to replace missing values of the dates of time of onset of symptoms with possible dates of infection in seropositive individuals without acute respiratory illness. Total of 1000 datasets were created where for each individual without acute respiratory illness the date was imputed from the dataset of confirmed COVID-19 cases [2] within the same county and in the inhabitants of the same age group detected until 14 days prior to drawing the individual’s blood sample. Coefficients of multiple linear regression models and their standard errors were calculated by Rubin’s rules [15].

## Results

### Study population

In total 3608 subjects were invited to participate of whom 1960 (54.3%) agreed and provided blood samples (Figure 1); all except one filled in the questionnaire. Of the participants 809 (41.3%) were male, 182 (9.3%) had been in contact with confirmed COVID-19 case, 221 (11.3%) had had PCR test for SARS-CoV-2 and 157 (8.0%) had had acute respiratory illness (Table 1). During the study period, the spread of SARS-CoV-2 had largely ended in Saaremaa, while in Tallinn it was ongoing (Figure S1).

**Table 1.** Seroprevalence and confidence interval (CI) in the two regions by general characteristics

|  | Saaremaa |  |  | Tallinn |  |
| --- | --- | --- | --- | --- | --- |
|  | Number of participants | Seroprevalence, %<br>(95% CI) |  | Number of participants | Seroprevalence, %<br>(95% CI) |
| **Overall** | 954 | 6.3<br>(5.0-7.9) |  | 1006 | 1.5<br>(0.9-2.5) |
| **Sex** |  |  |  |  |  |
| Male | 365 | 6.5<br>(4.4-9.4) |  | 444 | 0.5<br>(0.1-1.8) |
| Female | 589 | 6.1<br>(4.5-8.3) |  | 562 | 2.3<br>(1.3-4.1) |
| **Age group (years)** |  |  |  |  |  |
| 0-9 | 101 | 5.9<br>(2.2-12.5) |  | 106 | 3.8<br>(1-9.4) |
| 10-19 | 110 | 5.5<br>(2-11.5) |  | 107 | 0<br>(0-3.4) |
| 20-29 | 104 | 5.8<br>(2.1-12.1) |  | 111 | 0.9<br>(0-4.9) |
| 30-39 | 121 | 7.4<br>(3.5-13.7) |  | 117 | 0.9<br>(0-4.7) |
| 40-49 | 114 | 7<br>(3.1-13.4) |  | 120 | 1.7<br>(0.2-5.9) |
| 50-59 | 113 | 7.1<br>(3.1-13.5) |  | 117 | 1.7<br>(0.2-6) |
| 60-69 | 112 | 5.4<br>(2-11.3) |  | 116 | 2.6<br>(0.5-7.4) |
| 70-79 | 112 | 3.6<br>(1-8.9) |  | 114 | 0<br>(0-3.2) |
| 80-100 | 67 | 9<br>(3.4-18.5) |  | 98 | 1<br>(0-5.6) |
| **Acute respiratory illness** |  |  |  |  |  |
| Yes | 85 | 15.6<br>(9.6-24.4) |  | 72 | 1.7<br>(0.3-10.8) |
| No | 869 | 5.3<br>(4.1-6.9) |  | 933 | 1.4<br>(0.8-2.5) |
| **PCR** |  |  |  |  |  |
| Tested | 135 | 15.1<br>(10.2-21.8) |  | 86 | 10.7<br>(5.3-20.5) |
| PCR-negative | 117 | 1.3<br>(0.3-5.1) |  | 81 | 5.1<br>(1.4-16.3) |
| Not tested | 819 | 4.9<br>(3.6-6.5) |  | 919 | 0.6<br>(0.3-1.3) |
| **Household size** |  |  |  |  |  |
| 1 | 14 | 0.0<br>(0-23.1)* |  | 15 | 6.3<br>(0.9-32.1) |
| 2 | 157 | 6.4<br>(3.6-11.0) |  | 189 | 1.5<br>(0.4-6.0) |
| 3 | 268 | 6.9<br>(4.5-10.4) |  | 285 | 0.4<br>(0.1-1.6) |
| 4 | 190 | 6.6<br>(4.0-10.8) |  | 214 | 2.2<br>(0.9-5.1) |
| 5 | 185 | 5.6<br>(3.1-9.8) |  | 219 | 2.0<br>(0.8-5.2) |
| >5 | 140 | 6.2<br>(3.3-11.5) |  | 83 | 0.0<br>(0-4.3)* |
| **Contact with confirmed case** |  |  |  |  |  |
| Yes | 163 | 16.9<br>(12.2-23.0) |  | 19 | 24.2<br>(9.7-48.6) |
| No | 791 | 4.0<br>(2.9-5.5) |  | 986 | 0.9<br>(0.5-1.8) |

\*95% CI calculated for raw seroprevalence estimates.

### Laboratory testing

In total, 73 participants were seropositive. Of those all had detectable concentrations of anti-RBD by LIPS assay, 71 (97.3%) anti-N IgG by Abbott or anti-S1/S2 by DiaSorin assay, 68 (93.2%) anti-N IgG by LIPS assay, 60 (82.2%) neutralizing antibodies, 44 (60.3%) anti-N IgG by Biosensor assay (Figure 2). All 146 samples from seronegative age- and sex-matched controls were negative for anti-N IgG by Biosensor assay; of 22 samples tested by LIPS assays, one was positive for anti-N IgG.

**Figure 2.**
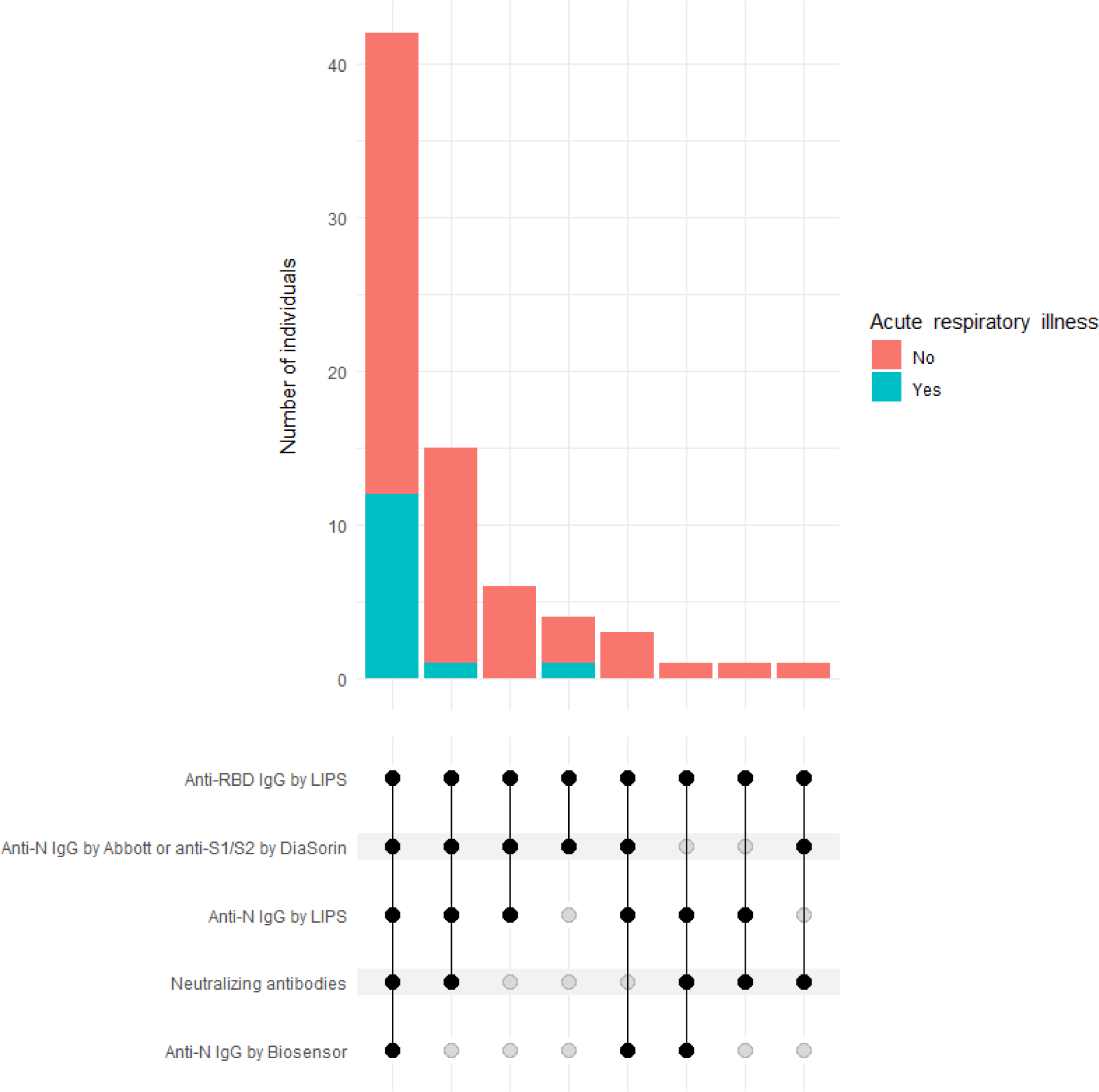
Upset plot of number of seropositive individuals with positive test results in different assays. Each bar shows number of individuals with positive results from the assays indicated by black circles beside the plot. Bars are colored according to number of individuals who had had acute respiratory illness (red) or not (green). Circles in the bottom show for each set of individuals negative (transparent) or positive (black) test result by the respective assay. Solid lines connect black circles.

### Seroprevalence

Overall seroprevalence was 1.5% (95% CI 0.9-2.5) in Tallinn and 6.3% (95% CI 5.0-7.9) in Saaremaa (Table 1). There were no difference in seroprevalence between males and females, age groups or household sizes. Seroprevalence was higher in the participants with contact with confirmed case or acute respiratory illness compared with those without. The seroprevalence estimates adjusted for test performance were similar to the estimates not adjusted for test performance, both overall (1.5% (95% CI 0.6-2.4) in Tallinn, 6.7% (95% CI 5-8.4) in Saaremaa) and in age groups (Table S1).

Number of deaths due to COVID-19 was 7 in Tallinn and 26 in Saaremaa and the respective infection fatality rates were 0.1% (95% CI 0.0-0.2) and 1.3% (95% CI 0.4-2.1).

### Seroprevalence vs confirmed cases

The cumulative number of confirmed COVID-19 cases in Tallinn (as of June 10, 2020) was 11.3 and in Saaremaa (as of June 15, 2020) 167.2 per 10,000 inhabitants. Seroprevalence of COVID-19 in Tallinn was 13.3 (range 8.0-22.1 based on 95% CI of seroprevalence estimate) and in Saaremaa 3.8 (3.0-4.7) times greater than the cumulative incidence of COVID-19 reported in the respective regions.

Of individuals with contact with confirmed case 43.0% (95/221) and of those with acute respiratory illness 33.1% (52/57) had PCR test performed. Considering participants without PCR test performed, in those with contact with confirmed case the proportion of seropositive individuals was 13.5% (17/126) and in those without contact 1.5 % (29/1612) (p<0.001); in those with acute respiratory illness the proportion of seropositive individuals was 6.7% (7/105) and in those without illness 2.4% (39/1633) (p=0.018).

### Symptoms

Of seropositive individuals 19.2% (14/73) and of seronegative individuals 7.6% (143/1886) self-reported having had acute respiratory illness (p=0.001) with median (interquartile range) of 6 (5-7) and 5 (3-7) symptoms, respectively. The frequency of different symptoms was similar in seropositive and seronegative individuals with acute respiratory illness, but seropositive individuals required more often hospitalization due to the acute respiratory illness compared with seronegative individuals (Table 2).

**Table 2.** Characteristics of acute respiratory illness since March 1, 2020 in seropositive or seronegative individuals

|  | Seropositive<br>(n=14) | Seronegative (n=143) |
| --- | --- | --- |
| Fever $\geq 38^{\circ}\text{C}$ – n (%) | 7 (50) | 45 (31.5) |
| Chills – n (%) | 7 (50) | 57 (39.9) |
| Fatigue – n (%) | 11 (78.6) | 96 (67.1) |
| Muscle aches – n (%) | 7 (50) | 45 (31.5) |
| Sore throat – n (%) | 5 (35.7) | 88 (61.5) |
| Cough – n (%) | 6 (42.9) | 100 (69.9) |
| Runny nose – n (%) | 8 (57.1) | 99 (69.2) |
| Difficulty breathing – n (%) | 5 (35.6) | 34 (23.8) |
| Shortness of breath – n (%) | 4 (28.6) | 22 (15.4) |
| Chest pain – n (%) | 4 (28.6) | 29 (20.3) |
| Other respiratory symptoms – n (%) | 2 (14.3) | 21 (14.7) |
| Headache – n (%) | 10 (71.4) | 75 (52.4) |
| Nausea/vomiting – n (%) | 0 | 17 (11.9) |
| Stomach ache – n (%) | 2 (14.3) | 19 (13.3) |
| Diarrhea – n (%) | 5 (35.7) | 32 (22.4) |
| Sought medical attention – n (%) | 8 (57.1) | 90 (62.9) |
| Missed work or school – n (%) | 8 (57.1) | 61 (42.7) |
| Hospitalized* – n (%) | 3 (21.4) | 7 (4.9) |

\*Only difference between the proportion of hospitalized patients between seropositive and seronegative individuals was statistically significant ( $p=0.047$ ).

In MCA of all individuals with acute respiratory illness, the first two dimensions discriminated poorly seropositive and seronegative subjects (Figure S2). Seropositivity had high quality on dimensions three and four and was associated with the absence of cough and the absence of sore throat (Figure S2). In MCA of 102 participants aged less than 50 years, of whom 5 were seropositive, it was not possible to associate seropositivity with any symptom by the first 6 dimensions due to low quality and contribution of seropositivity on dimensions 1-4 or symptoms on 5-6. In MCA of 55 participants aged 50 years or more, of whom 9 were seropositive, according to the first two dimensions of MCA seropositivity was associated with the absence of cough and runny nose and the presence of diarrhea and fever (Figure 3) and majority (7/9) of seropositive individuals were grouped into a single k-means cluster containing no seronegative individuals (Figure S3).

**Figure 3.**
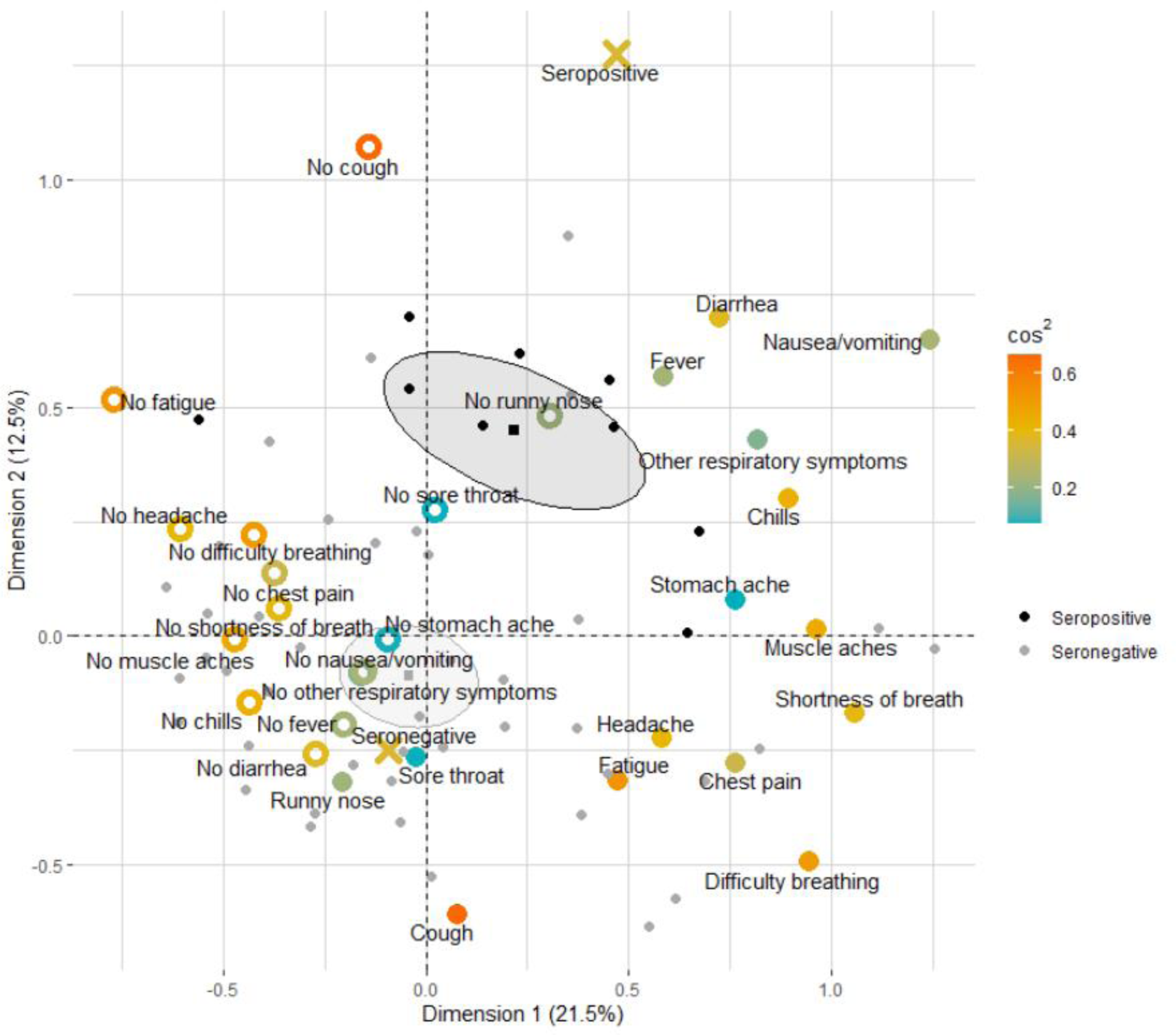
Biplot of the first two dimensions of multiple correspondence analysis on association between the presence (filled large dots) or absence (empty large dots) of symptoms and seropositivity (crosses) in all individuals aged 50 years or more with acute respiratory illness. The variables included in the analysis are colored according to the sum of cos^2^ of the variable on dimensions one and two. Grey and black small dots represent seronegative and seropositive individuals, respectively. Practices of gravity of seropositive and seronegative individuals are shown by squares surrounded by 95% confidence ellipses.

### Antibody concentrations/titer

According to multiple linear regression, anti-N IgG and anti-RBD IgG concentrations by LIPS assay and anti-N IgG by Abbott assay were significantly higher in seropositive individuals with fever (Table 3). Anti-RBD and anti-N IgG by LIPS assay were significantly higher in hospitalized patients, anti-N IgG by LIPS and/or Abbott assay in patients with difficulty breathing, shortness of breath or chest pain and anti-N IgG by Abbott assay in patients with diarrhea. There were no statistically significant associations between titer of neutralizing antibodies and characteristics of acute respiratory illness.

**Table 3.** Regression coefficients and 95% confidence intervals of multiple linear regression models of association between characteristics of acute respiratory illness and IgG concentrations or neutralizing antibody titer in seropositive individuals, adjusted for age, sex or time since onset of symptoms or multiply imputed positive PCR test

|  | Anti-RBD IgG by LIPS | Anti-N IgG by LIPS | Anti-N IgG by Abbott | Neutralizing antibodies |
| --- | --- | --- | --- | --- |
| Acute respiratory illness | 29.0 (-19.0...77.0) | 5.0 (-0.1...10.1) | 1.5 (-0.1...3.0) | 0.6 (-0.7...1.9) |
| Fever $\geq 38^{\circ}\text{C}$ | **62.4**<br>**(0.6...124.2)\*** | **9.8**<br>**(3.4...16.3)\*\*** | **3.0**<br>**(1.1...4.9)\*\*** | 1.2 (-0.5...2.9) |
| Chills | -10.5 (-73.8...52.8) | 1.6 (-5.2...8.4) | 0.4 (-1.6...2.5) | 0.01 (-1.7...1.7) |
| Fatigue | 14.2 (-37.0...65.5) | 4.5 (-0.9...10.0) | 1.1 (-0.6...2.7) | 0.3 (-1.0...1.7) |
| Muscle aches | 23.3 (-37.0...83.5) | 4.2 (-2.3...10.7) | 1.3 (-0.6...3.2) | 0.5 (-1.1...2.2) |
| Sore throat | -50.1<br>(118.9...18.7) | -2.9 (-10.4...4.6) | 0.3 (-2.0...2.5) | -0.5 (-2.4...1.3) |
| Cough | 20.9 (-45.3...87.1) | 5.6 (-1.5...12.6) | 1.7 (-0.4...3.8) | 0.6 (-1.2...4.1) |
| Runny nose | 12.3 (-46.4...71.0) | 3.8 (-2.5...10.0) | 1.5 (-0.3...3.4) | 1.0 (-0.5...2.6) |
| Difficulty breathing | 69.7 (-4.1...143.5) | **9.3**<br>**(1.4...17.1)\*** | 2.3 (-0.1...4.6) | 1.0 (-1.0...3.1) |
| Shortness of breath | 53.6 (-25.8...132.9) | **9.4**<br>**(1.1...17.8)\*** | **2.6 (0.1...5.1)\*** | 1.7 (-0.4...3.9) |
| Chest pain | 46.0 (-31.2...123.3) | 8.1 (-0.1...16.4) | **3.2**<br>**(0.9...5.6)\*\*** | 1.0 (-1.1...3.1) |
| Other respiratory symptoms | -8.9 (-116.2...98.4) | 2.7 (-8.8...14.3) | 1.5 (-1.9...5.0) | -0.1 (-3.0...2.8) |
| Headache | 4.1 (-49.2...57.4) | 1.9 (-3.8...7.6) | 0.5 (-1.2...2.2) | 0.1 (-1.3...1.6) |
| Stomach ache | -60.2 (-165.4...45.0) | -7.6 (-18.9...3.7) | -1.0 (-4.4...2.4) | -2.0 (-4.8...0.9) |
| Diarrhea | 31.1 (-38.2...100.4) | 6.4 (-1.0...13.7) | **2.5 (0.4...4.7)\*** | 0.3 (-1.6...2.2) |
| Hospitalized | **102.8**<br>**(13.6...192.0)\*** | **10.5**<br>**(0.9...20.2)\*** | 2.7 (-0.2...5.6) | 1.3 (-1.2...3.8) |

Statistically significant regression coefficients are shown in bold.
\* p-value between 0.01 to 0.05, \*\* p-value <0.01.

## Discussion

To the best of our knowledge this is the first seroepidemiological study from Eastern Europe with strengths of including all age groups and using several assays for antibody testing. Our study showed that the seroprevalence of COVID-19 in two regions of Estonia with very different extent of the spread of SARS-CoV-2 by PCR in March and April 2020 were 1.5% and 6.3% with no difference between age groups. Most seropositive individuals had not had acute respiratory illness and only 4% were hospitalized. In individuals aged 50 years or more fever, diarrhea and the absence of cough and runny nose were associated with seropositivity. The concentrations of IgG, but not titer of neutralizing antibodies, were higher if fever, or difficulty breathing, shortness of breath, chest pain or diarrhea was present, or hospitalization was required.

### Seroprevalence

The seroprevalence estimates found in our study are within the range of 0.4-10.9% reported in other population-based seroepidemiological studies [4-8] and far from the threshold of 60 to 70% required for herd immunity [16]. Considering the number of cumulative cases per 10,000 inhabitants of 167.2, we would have expected larger seroprevalence in Saaremaa, as seroprevalence estimates were between 4.6-10.9% in Spain, United Kingdom and Geneva [4, 6, 7] with smaller number of cumulative cases per 10,000 (44.7-103.2) and thereby larger infection-to-case ratios 10.6-14.3 [1, 4]. However, these seroepidemiological studies were conducted shortly after the peak of the spread when the daily incidence curve had flattened at low levels, but the spread was still ongoing [1, 4, 6, 7], suggesting the presence of undocumented cases and thereby underestimation of cumulative cases per 10,000 inhabitants. Similar pattern of ongoing spread during sample collection for seroepidemiological study and infection-to-case ratio 11.3 was in Tallinn. In contrast, in Saaremaa the spread of SARS-CoV-2 had been successfully contained and only few cases were detected during the study period, a situation similar to Reykjavik and Luxembourg, where the cumulative cases per 10,000 inhabitants were 50.4 and 53.3 and seroprevalence estimates 0.4% and 2.0%, respectively [5, 8]. Such discrepancies between infection-to-case ratios underline once again the importance of seroepidemiological studies in estimating the actual prevalence of infection. It is believed that incidence of SARS-CoV-2 infection was severely underreported in spring not only in Estonia but elsewhere [17]. This is further supported by the age distribution of confirmed cases with predominance of older age groups (46.1% and 68.4% aged 50 years or more in Tallinn and Saaremaa, respectively [2]) among reported cases while age distribution in our study was even in our seroepidemiological study.

### Symptoms

Of individuals with acute respiratory illness, two thirds had not been tested by PCR, but they were significantly more likely to be seropositive, showing underdetection of cases among them. Detection of cases by individual symptoms has low specificity [18, 19], particularly during the season of respiratory infections [18], but this could be increased by a combination of them [20]. Small number of individuals with acute respiratory illness impeded detection of pattern in our study. Although the proportion of asymptomatic cases among PCR-positive individuals has been up to 87.9% [21], we may have missed some symptomatic cases due to focusing only on acute respiratory illness. However, the proportion of COVID-19 cases presenting without any respiratory symptoms, such as solely by fever or gastrointestinal symptoms, is low, up to 21% [22]. Furthermore, it is notable that while the proportion of fever, the most specific symptom of COVID-19 [18], in cases detected by PCR is larger than that of cough [18, 23], in seropositive individuals fever is mostly less common or occurs with rate comparable to cough [8, 24]. This may partly corroborate that the proportion of symptoms in seropositive cases attributable causes other than COVID-19 may be notable, up to 31.5% [25]. Additionally, this may suggest underdetection of cases among individuals with respiratory infections presenting without fever, supported by smaller proportion of people with cough being tested compared to those with fever [26], warranting focusing particularly on respiratory infections, as in our study. In adults aged 50 years or more, we found that the symptoms associated with seropositivity were the absence of runny nose and cough and the presence fever and diarrhea, similarly to a previous study [27], but the lack of information on anosmia or ageusia that are most strongly associated with COVID-19 [24, 26] may have compromised our analysis. Nevertheless, our results suggest that focusing only on respiratory symptoms may delay accurate diagnosis of SARS-CoV-2 infection in older people.

### Antibody concentrations/titer

In line with several other studies, we found that hospitalized patients or patients with symptoms suggesting more severe disease, such as fever, or shortness of breath had higher anti-N IgG concentrations [5, 14, 28]. However, these factors were not associated with titer of neutralizing antibodies or were less often associated with anti-RBD IgG that contain large proportion of neutralizing antibodies [29], in contrast with other studies [14, 30]. While these studies are mostly conducted on samples collected within two months after development of symptoms, long-term response of neutralizing antibodies is scarcely described [28]. Notably, in contrast to anti-RBD and anti-N IgG, neutralizing antibodies decline to undetectable titer two months after the onset of symptoms in 20-25%of patients, as in our study, despite development of detectable peak in nearly all of them [28]. Moreover, titers of neutralizing antibodies decline faster compared with anti-RBD and anti-N IgG [14, 28]. Therefore, no associations found for neutralizing antibodies could be attributable to such faster decline and possible attainment of a plateau concentration where differences are less clear, as suggested [28], but further studies on longevity of neutralizing antibodies are warranted.

## Limitations

Our study has some limitations. First, as only 54% of individuals asked to participate in the study were willing to do so, participation bias may affect the results of our study. Second, as we did not sample from population register of Saaremaa and Tallinn, but from the list of patients of GP practices, the sampled individuals and thus seroprevalence estimates may not be representative of the regions and not directly comparable to confirmed cases. However, the age distribution of GP practice patients resembled that of the respective counties and testing rates in people near the GP practices was similar to the overall county [2]. Third, our testing algorithm for detection of seropositive individuals has unknown specificity and sensitivity and thus we cannot accurately estimate seroprevalences adjusted for test performance. Furthermore, we cannot rule out exclusion of false-negative individuals by testing only with combination of Abbott and DiaSorin assay, but such approach increases sensitivity by 7% [10].

In conclusion, seroprevalence of COVID-19 in Estonia in spring 2020 was low despite large number of confirmed cases, underlining the importance of seroepidemiological studies. Most people with SARS-CoV-2 infections had mild disease or did not report any respiratory symptoms. Improving detection of cases among individuals with contact with confirmed cases or with acute respiratory illness or with fever and diarrhea in those aged 50 years or more may contribute to containment of the upsurge of the virus, but further long-term studies are warranted to demonstrate persistence of antibodies.

## Data Availability

Data of this study is available from the corresponding author, PJ, upon reasonable request.

## Acknowledgements

We thank prof Krista Fischer for her advice in statistical analysis and all the study participants. The study was funded by the Ministry of Social Affairs of the Republic of Estonia. Electronic data collection was supported by the High Performance Computing Center of the University of Tartu.

## Supplementary Content

### Supplementary Information 1

Study questionnaire

### 1. Personal data

|  |  |
| --- | --- |
| 1.1. | First name |
| 1.2. | Family name |
| 1.3. | Patient ID |
| 1.4. | Age (in years) |
| 1.5. | Phone number |
| 1.6. | E-mail |
| 1.7. | Place of residence (county) |
| 1.8. | Number of family members living together |
| 1.9. | Form completion date (dd/mm/yyyy) |

## 2. History of COVID-19

|  |  |  |
| --- | --- | --- |
|  | Yes | No |
| 2.1. Have you had acute respiratory illness since March 1, 2020? |  |  |

### 2.1.1 If Yes, then date of becoming ill? ......../........./............... (dd/mm/yyyy)

2.1.1. If Yes, then date of becoming ill? ...../...../..... (dd/mm/yyyy)
|  |  |  |
| --- | --- | --- |
|  | Yes | No |
| 2.2. Have you had a COVID-19 PCR test (nasal swab)? |  |  |
| 2.2.1. If Yes, then did the PCR test confirm COVID-19 infection? |  |  |
| 2.3. Have you been in contact with someone who has been diagnosed with COVID-19 infection? |  |  |

### 2.3.1 If Yes, then date of the last contact? ........./........../................ (dd/mm/yyyy)

2.3.1. If Yes, then date of the last contact? ...../...../..... (dd/mm/yyyy)
|  |  |  |  |
| --- | --- | --- | --- |
|  | Yes | No | Not sure |
| 2.4. Have you been vaccinated against tuberculosis in childhood? |  |  |  |

## 3. Symptom history (only for those who answered Yes in 2.1)

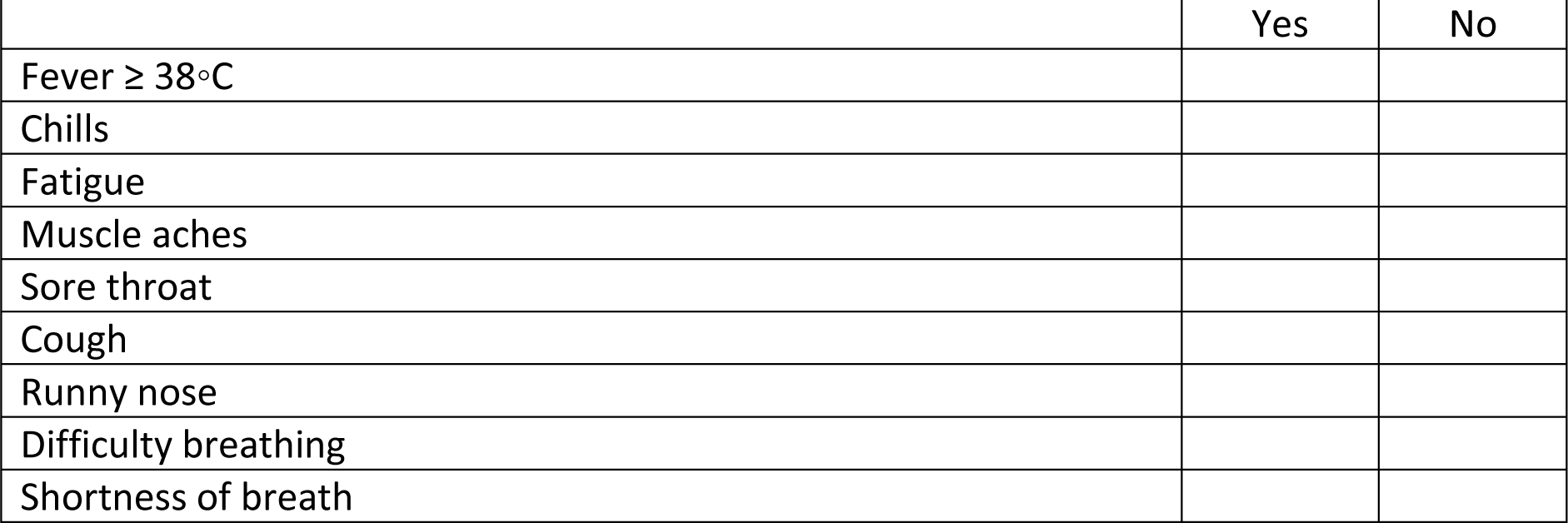
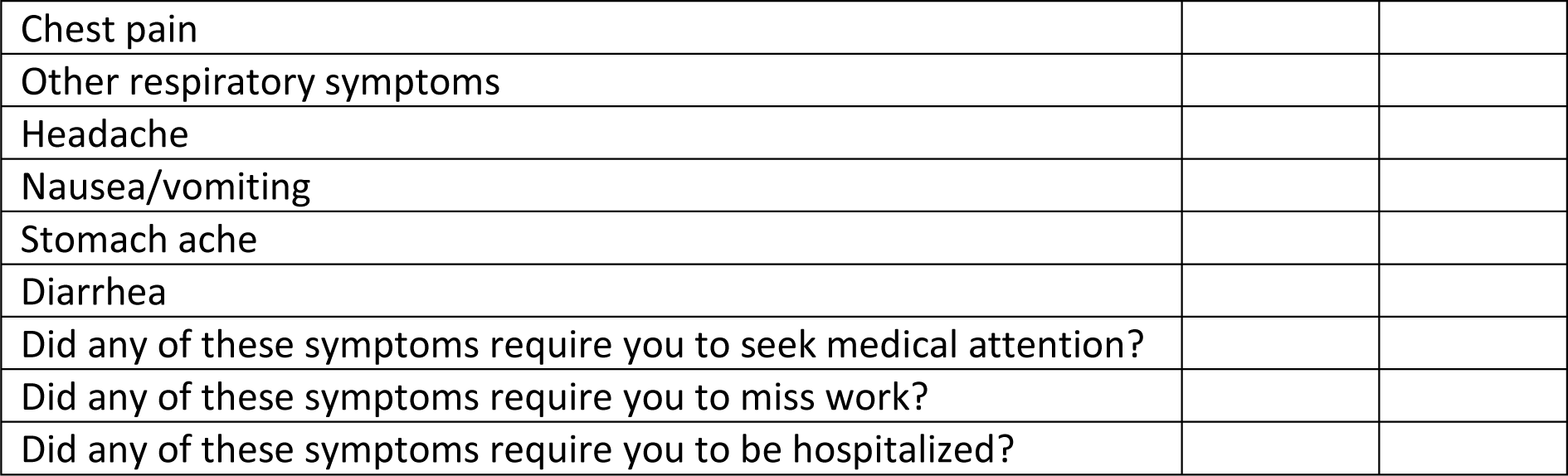

|  |  |  |
| --- | --- | --- |
|  | Yes | No |
| Fever $\geq 38^{\circ}\text{C}$ | | |
| Chills |  |  |
| Fatigue |  |  |
| Muscle aches |  |  |
| Sore throat |  |  |
| Cough |  |  |
| Runny nose |  |  |
| Difficulty breathing |  |  |
| Shortness of breath |  |  |

|  |
| --- |
| Chest pain |
| Other respiratory symptoms |
| Headache |
| Nausea/vomiting |
| Stomach ache |
| Diarrhea |
| Did any of these symptoms require you to seek medical attention? |
| Did any of these symptoms require you to miss work? |
| Did any of these symptoms require you to be hospitalized? |

**Table S1.** Seroprevalence and seroprevalence adjusted for test performance according to age groups and confidence interval (CI)

| Age group (years) | Seroprevalence,<br>% (95% CI) | Seroprevalence adjusted for test<br>performance, % (95% CI) |
| --- | --- | --- |
| **Tallinn** |  |  |
| 0-9 | 3.8 (1-9.4) | 4.0 (0.8-8.1) |
| 10-19 | 0 (0-3.4) | 0.0 (0-0) |
| 20-29 | 0.9 (0-4.9) | 0.9 (0-2.9) |
| 30-39 | 0.9 (0-4.7) | 0.8 (0-2.8) |
| 40-49 | 1.7 (0.2-5.9) | 1.7 (0-4.5) |
| 50-59 | 1.7 (0.2-6) | 1.7 (0-4.6) |
| 60-69 | 2.6 (0.5-7.4) | 2.7 (0-6.3) |
| 70-79 | 0 (0-3.2) | 0.0 (0-0) |
| 80-100 | 1 (0-5.6) | 1.0 (0-3.3) |
| **Saaremaa** |  |  |
| 0-9 | 5.9 (2.2-12.5) | 6.3 (2-11.7) |
| 10-19 | 5.5 (2-11.5) | 5.8 (1.8-10.7) |
| 20-29 | 5.8 (2.1-12.1) | 6.1 (1.9-11.4) |
| 30-39 | 7.4 (3.5-13.7) | 7.9 (3.4-13.4) |
| 40-49 | 7 (3.1-13.4) | 7.5 (2.7-13) |
| 50-59 | 7.1 (3.1-13.5) | 7.5 (2.8-13.1) |
| 60-69 | 5.4 (2-11.3) | 5.7 (1.7-10.6) |
| 70-79 | 3.6 (1-8.9) | 3.7 (0.7-7.8) |
| 80-100 | 9 (3.4-18.5) | 9.6 (3.1-17.6) |

**Figure S1.**
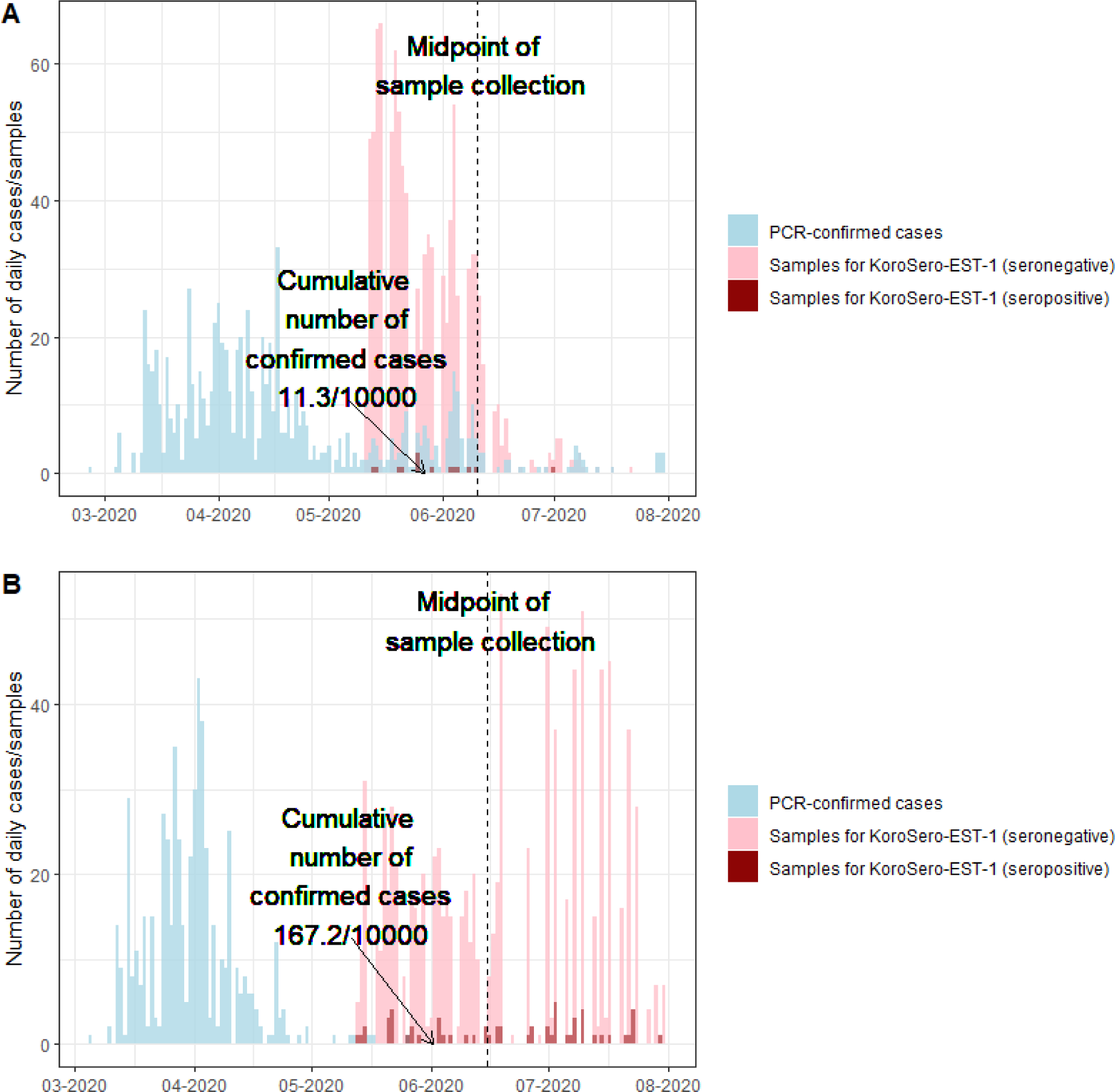
PCR-confirmed cases (blue) and number of seropositive (dark red) or seronegative (pink) serum samples drawn for the seroepidemiological study KoroSero-EST-1 (A) in Tallinn and (B) in Saaremaa

**Figure S2.**
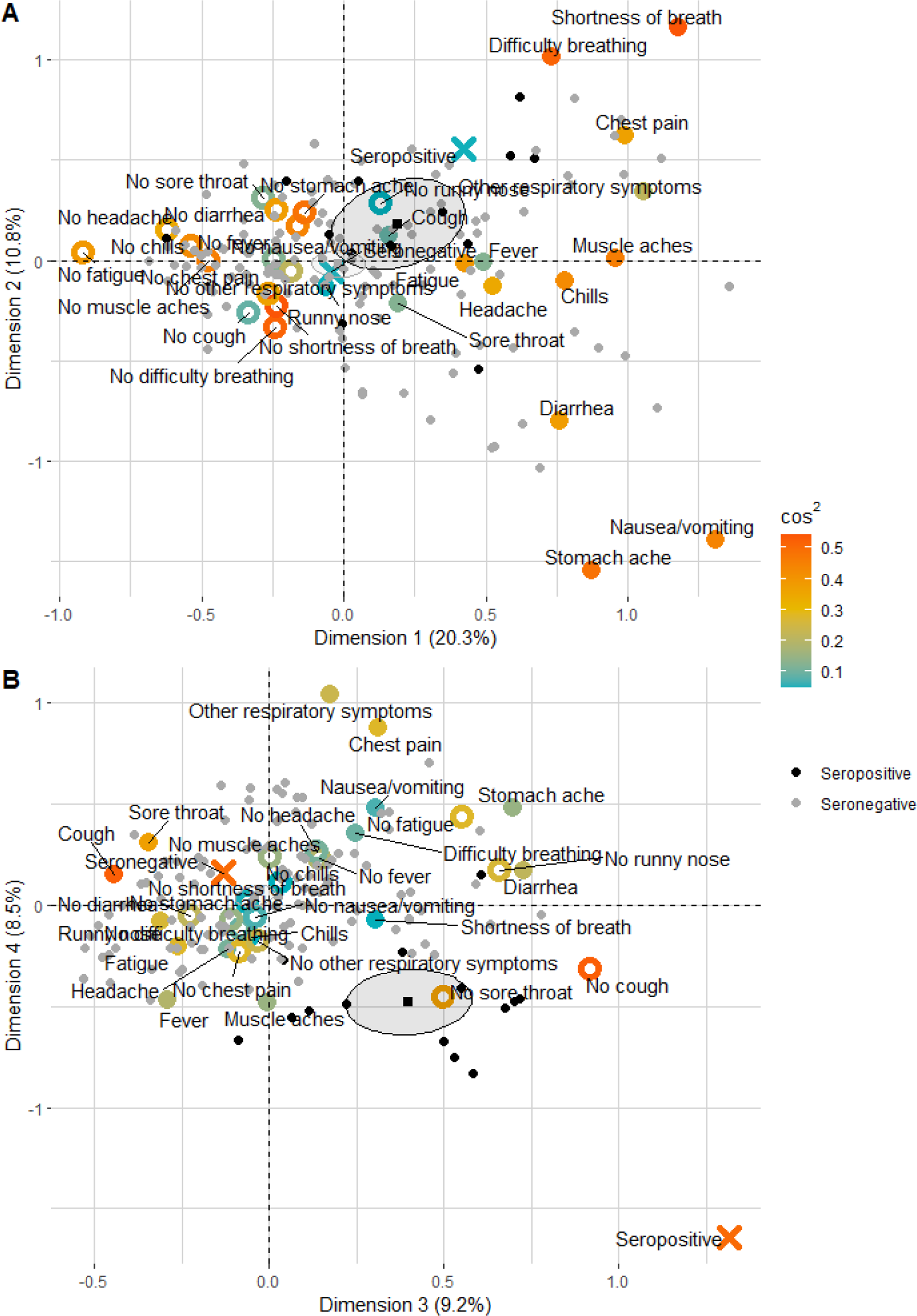
Biplot of the dimensions 1 and 2 (panel A) and 3 and 4 (panel B) of multiple correspondence analysis on association between the presence (filled large dots) or absence (empty large dots) of symptoms and seropositivity (crosses) in all individuals with acute respiratory illness. The variables included in the analysis are colored according to the sum of cos^2^ of the variable on dimensions 1 and 2 (panel A) or 3 and 4 (panel B). Grey and black small dots represent seronegative and seropositive individuals, respectively. Practices of gravity of seropositive and seronegative patients are shown by squares surrounded by 95% confidence ellipses. According to dimensions three and four seropositivity was associated with the absence of cough and the absence of sore throat

**Figure S3.**
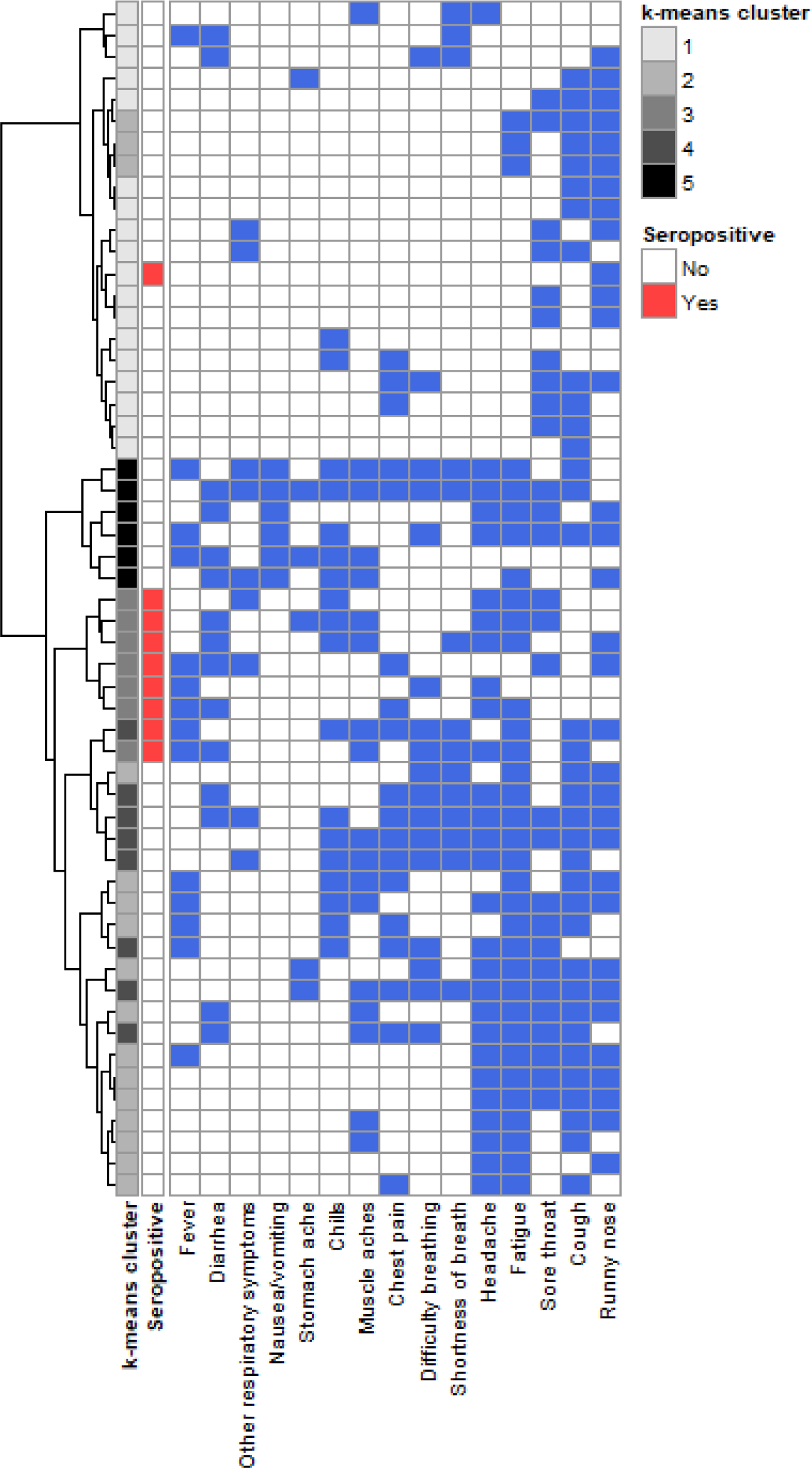
Clustering of individuals aged 50 years or more with acute respiratory illness according to hierarchical clustering on components of multiple correspondence analysis and subsequent k-means consolidation into 5 groups (leftmost column, designated with different shades of grey). Seropositivity is shown in the second leftmost column in red. The presence of the respective symptom is indicated in blue. The majority (7/9) of seropositive individuals are grouped into a single k-means cluster containing no seronegative individuals.

